# Immunopathological characteristics of coronavirus disease 2019 cases in Guangzhou, China

**DOI:** 10.1101/2020.03.12.20034736

**Authors:** Yaling Shi, Mingkai Tan, Xing Chen, Yanxia Liu, Jide Huang, Jingyi Ou, Xilong Deng

## Abstract

Coronavirus disease 2019 (COVID-19) is a respiratory disorder caused by the highly contagious SARS-CoV-2. The immunopathological characteristics of COVID-19 patients, either systemic or local, have not been thoroughly studied. In the present study, we analyzed both the changes in the cellularity of various immune cell types as well as cytokines important for immune reactions and inflammation. Our data indicate that patients with severe COVID-19 exhibited an overall decline of lymphocytes including CD4+ and CD8+ T cells, B cells, and NK cells. The number of immunosuppressive regulatory T cells was moderately increased in patients with mild COVID-19. IL-2, IL-6, and IL-10 were remarkably up-regulated in patients with severe COVID-19. The levels of IL-2 and IL-6 relative to the length of hospital stay underwent a similar “rise-decline”pattern, probably reflecting the therapeutic effect. In conclusion, our study shows that the comprehensive decrease of lymphocytes, and the elevation of IL-2 and IL-6 are reliable indicators of severe COVID-19.

## Introduction

Coronavirus disease 2019 (COVID-19) is a respiratory disorder caused by a new coronavirus termed SARS-CoV-2^1^. This virus probably arose from an unidentified animal source and is subsequently transmitted from person to person^2^. Transmission of COVID-19 is through the air by coughing and sneezing, close personal contact, or touching a virus-contaminated object and then touching mouth, nose, or possibly eyes. COVID-19’s initial manifestations include cough, fever, and respiratory distress. The thorough clinical profile of COVID-19 has not been completely comprehended. Reported illnesses range from mild to severe or even death. The severity of COVID-19 varies among individual patients during the same outbreak and among patient groups during different outbreaks in different regions.

The immune response in COVID-19 patients, either systemic or local, has not been well studied even one month after the COVID-19 outbreak in China. Although the chest CT scan has suggested progressive pneumonia in COVID-19 patients, the inflammatory process and immune reaction have not been detailed. In one report, elevation of neutrophil cellularity, serum IL-6, and c-reactive protein was found together with lymphopenia in COVID-19 patients^3^. Other clinical studies indicate that COVID-19 patients develop lymphopenia and high-levels of various cytokines such as G-CSF, IP-10, MCP-1, MIP-1A, and TNF-α^4, 5^. The surge of proinflammatory cytokines incurred the cytokine storm which might induce viral sepsis and tissue/organ damages to result in shock or even multiple organ failure. SARS-CoV-2-specific antibodies seem to be produced in COVID-19 patients, suggesting the mounting of humoral responses^3^. However, the comprehensive status of either innate immunity or adaptive immunity in COVID-19 patients remains largely unknown.

In the current study, we analyzed multiple cytokines and immune cell populations in the blood of Chinese COVID-19 patients. Our study outlines the immunopathological profile in COVID-19 patients, and may provide helpful information for developing future therapies against SARS-CoV-2 infection.

## Materials and methods

### Patients

The study was approved by the Ethics Committee of Guangzhou Eighth People’s Hospital. Thirty-one individuals who were diagnosed to have mild/moderate COVID-19 symptoms (17 men, 14 women, average age=44.5), and 25 individuals who were diagnosed to show severe COVID-19 symptoms (18 males, 7 females, average age=66) in Guangzhou Eighth People’s Hospital between January 2020 and February 2020 were enrolled in the study with informed consent. Diagnosis of COVID-19 infection, pneumonia and clinical classification was based on the new coronavirus pneumonia diagnosis and treatment plan (trial version 6) issued by the National Health Committee of the People’s Republic of China. The clinical classifications are on the basis of the following manifestations: (1) mild/moderate: fever, respiratory tract symptoms, and pneumonia on chest CT scan. Respiratory rate > 30 beats/min, or mean oxygen saturation < 93% (2) severe: one of the following: respiratory distress with respiratory rate > 30 beats/min, peripheral capillary oxygen saturation ≤ 93%, the ratio of partial pressure of oxygen (PaO2) to fraction of inspired oxygen (FiO2) < 300, respiratory failure requiring mechanical ventilation, Shock, ICU admission required for combined organ failure, pulmonary pathological progression. All patients were confirmed by the test on upper respiratory throat swab samples using the standard COVID-19 test kit. Individuals with fever and negative for the SARS-CoV-2 test were recruited as controls.

### Sample collection

Two to four milliliters of peripheral venous blood was withdrawn from each patient into K3 EDTA blood collection tubes. The complete blood cell count was performed on an XN-A1 automatic blood analyzer (Sysmex). The reagents, calibrators, and quality controls were from the same vendor.

### Flow cytometry assay

To analyze serum cytokines, the whole blood was centrifuged at 500×g for 5 minutes at 4°C. The serum was carefully harvested and stored at −80°C before analysis. Multiple serum cytokines were quantified using the Human Th1/Th2 Cytokine Kit II (Guangzhou Weimi Bio-Tech) following the manufacturer’s manual.

To measure lymphocytes and T cell subsets, 100 µl of whole blood was incubated in 900 µl of Tris-NH_4_Cl buffer (Thermo Fisher Scientific) at room temperature for 5 minutes to lyse erythrocytes. After two washes with phosphate-buffered saline (PBS), cells were incubated with BD Multitest 6-Color TBNK Reagent or BD Multitest CD3/CD8/CD45/CD4 (BD Biosciences) following the vendor’s instructions. After another two washes with PBS, cells were resuspended in 500 µl of PBS

To evaluate regulatory T cells, erythrocytes were lysed as described above. The blood samples were then incubated with FITC-conjugated CD4 antibody, Violet 450-conjugated CD127 antibody, Percp-conjugated CD45 antibody, and APC-conjugated CD25 antibody (5µg/ml each, all from BD Biosciences) for 15 minutes on ice. After another two washes with PBS, cells were resuspended in 500 µl of PBS.

Samples were analyzed on a BD FACSCanto Plus flow cytometer. Among all collected events, single events were gated between FSC-A and FSC-H. Cell debris was excluded and intact cells were then gated from single events based on FSC-A and SSC. Each cell population was then detected based on the antibody staining.

### Statistics

The data were indicated as mean□±□standard deviation and measured by GraphPad Prism 6.0. Non parametric Kruskal-Wallis test or One-way ANOVA with post-hoc Tukey HSD test was applied to compare the mean values or mean ranks among different groups, depending on the presence or absence of Gaussian distribution.

## Results

### T cell cellularity is reduced in COVID-19 patients

To assess the general immune status of COVID-19 patients, we first quantified the absolute number of whole white blood cells (WBC) and immune cell populations in blood samples of patients at inpatient admission. As indicated in Figure 1A, WBC number was significantly decreased in mild patients as compared with the control group. However, WBC number was not remarkably changed in severe patients. Total lymphocyte number was decreased in severe patients but not in mild patients, suggesting the lymphopenia severe patients (Figure 1B). Neutrophils were increased in severe patients but not in mild patients (Figure 1C). The neutrophil-to-lymphocyte ratio (NLR), consistent with the change in lymphocytes, was moderately increased in mild patients and profoundly increased in severe patients, in comparison to the control group (Figure 1D). To analyze the adaptive immune cell populations, we conducted flow cytometry analysis on CD45+CD3+ total T cells, CD3+CD4+ T helper cells and CD3+CD8+ cytotoxic T cells (Figure 1E). We found a similar decrease in CD45+ lymphocytes in COVID-19 patients (Figure 1F). Total T cell number was significantly decreased in COVID-19 patients, but no significant difference was observed between mild and severe patients (Figure 1G). Similarly, CD4+ T helper cells and CD8+ cytotoxic T cells were diminished in COVID-19 patients as compared with the control group, but no significant difference was found between mild and severe patients (Figure 1H & 1I). The ratio between CD4+ T cells and CD8+ T cells was not changed (Figure 1J).

**Figure 1.**
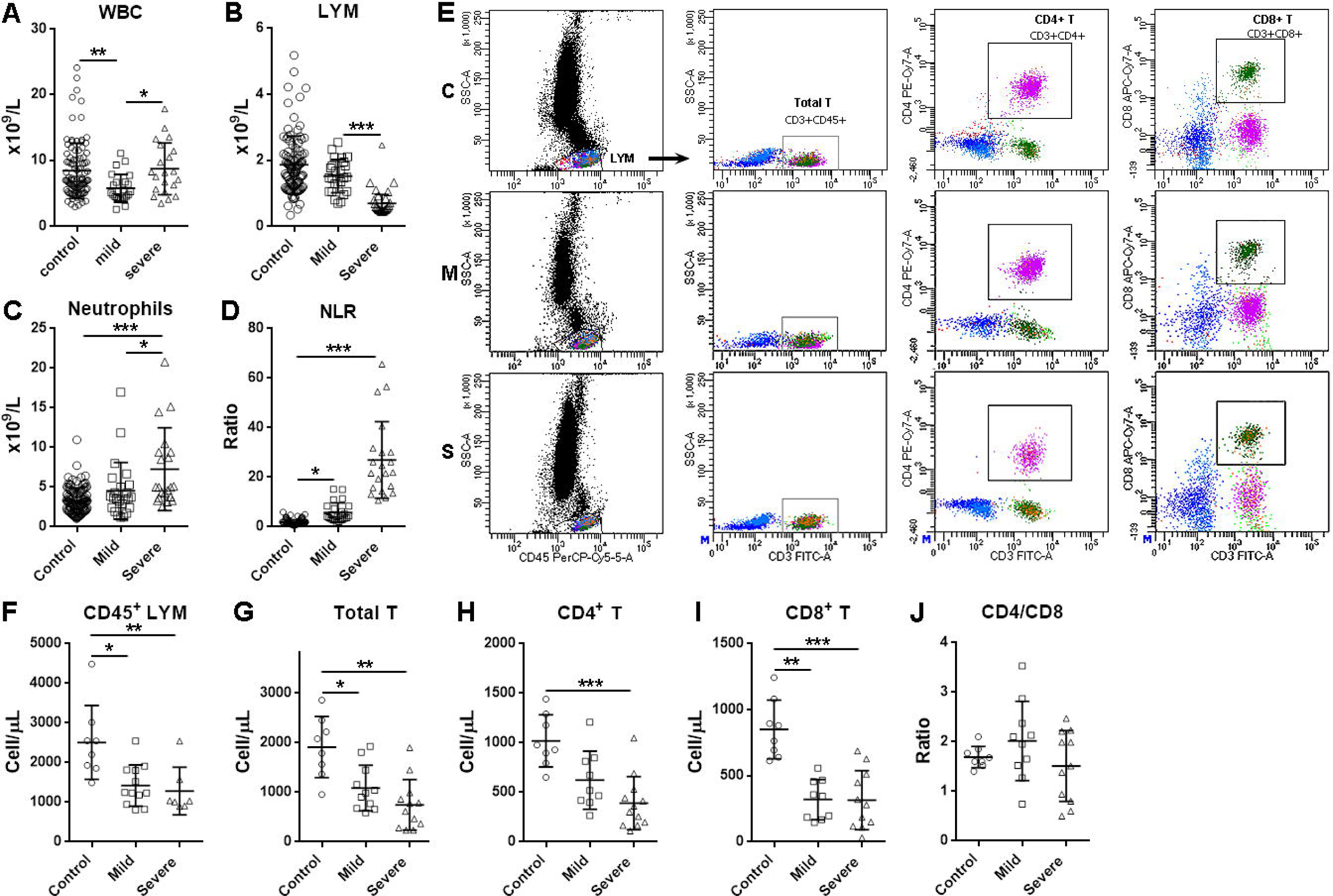
Cellularity of blood T cells in COVID-19 patients at hospital admission. **(A to C)** Absolute cell number of white blood cells (WBC), lymphocytes (LYM), and neutrophils. **(D)** Neutrophil-to-Lymphocyte ratios. **(E)** Representative flow cytometry dot plots showing the gating strategy for total T cells, CD4+ T cells, and CD8+ T cells. C: control; M: mild; S: severe. **(F to I)** Quantification of the number of CD45+ lymphocytes, total T cells, CD4+ T cells, and CD8+ T cells using flow cytometry analysis. **(J)** Ratio between CD4+ T cells and CD8+ T cells. *, p<0.05; **, p<0.01; ***, p<0.001.

### B cells and NK cells are decreased in COVID-19 patients

Other immune cells including CD3-CD19+ B cells, CD3-CD16+CD56+ NK cells and CD3+CD16+CD56+ NKT cells were also evaluated in blood samples (Figure 2A). We found that in comparison with the control group, there was a trend of decrease in B cell cellularity in mild COVID-19 patients, and a notable reduction in B cells of severe COVID-19 patients (Figure 2B). A similar decrease was also seen in COVID-19 patients’ NK cells (Figure 2C). No remarkable change in NKT cellularity was observed in COVID-19 patients (Figure 2D).

**Figure 2.**
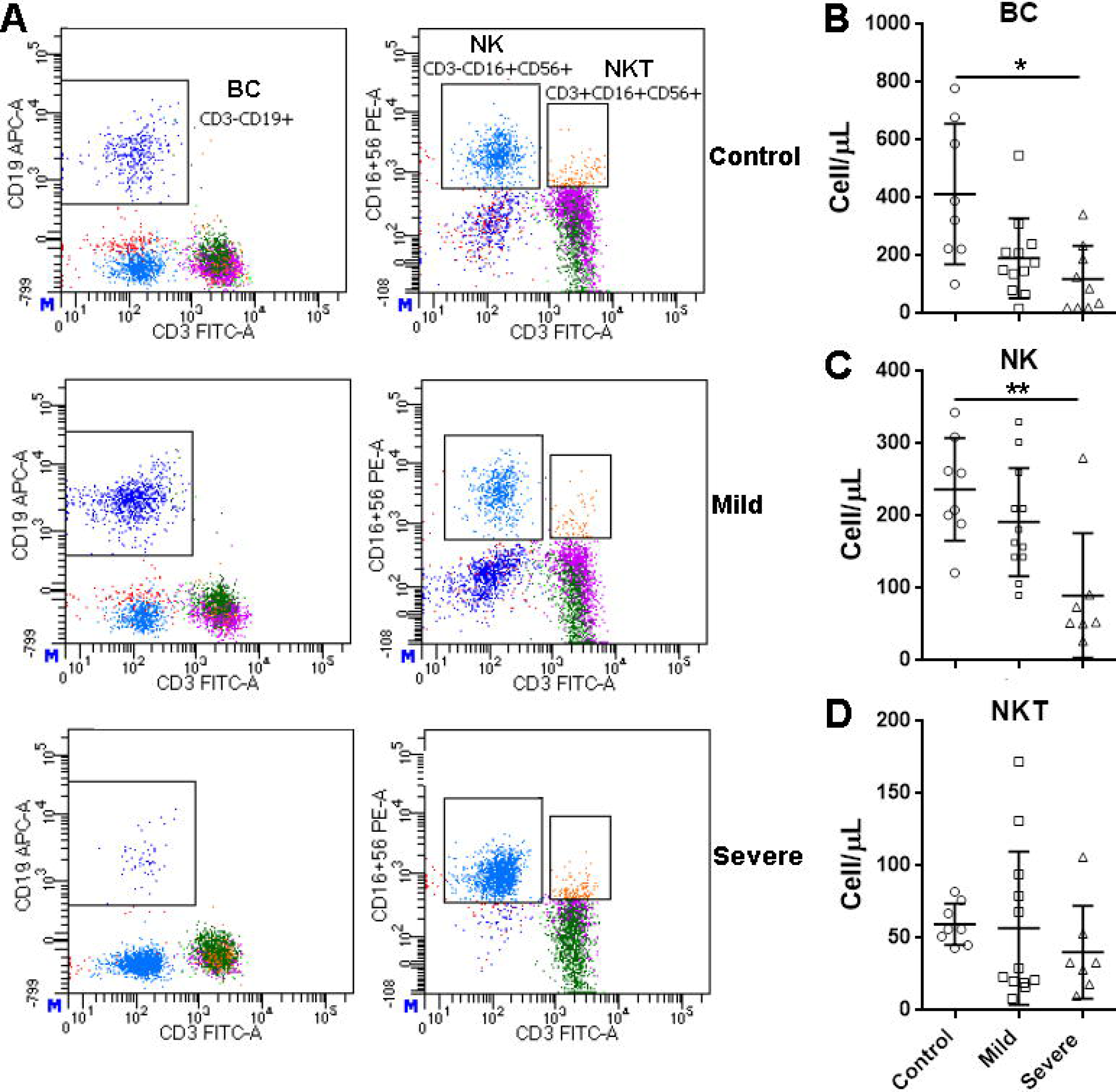
Cellularity of blood B cells, NK cells and NKT cells in COVID-19 patients at hospital admission. **(A)** Representative flow cytometry dot plots showing the gating strategy for B cells, NK cells and NKT cells. **(B to D)** Quantification of the number of B cells **(B)**, NK cells **(C)** and NKT **cells (D)**. *, p<0.05; **, p<0.01.

### Regulatory T cells (Tregs) are increased in mild COVID-19 patients

Tregs are an important T cell subset for immune tolerance and anti-inflammatory reaction. In our study, we measured the cellularity of CD3+CD4+CD25+CD127-T cells which were considered a Treg-enriched population (Figure 3A). We found that Tregs were increased in mild COVID-19 patients as compared with controls (Figure 3B). There was only a slight increase in Tregs in severe patients (Figure 3B).

**Figure 3.**
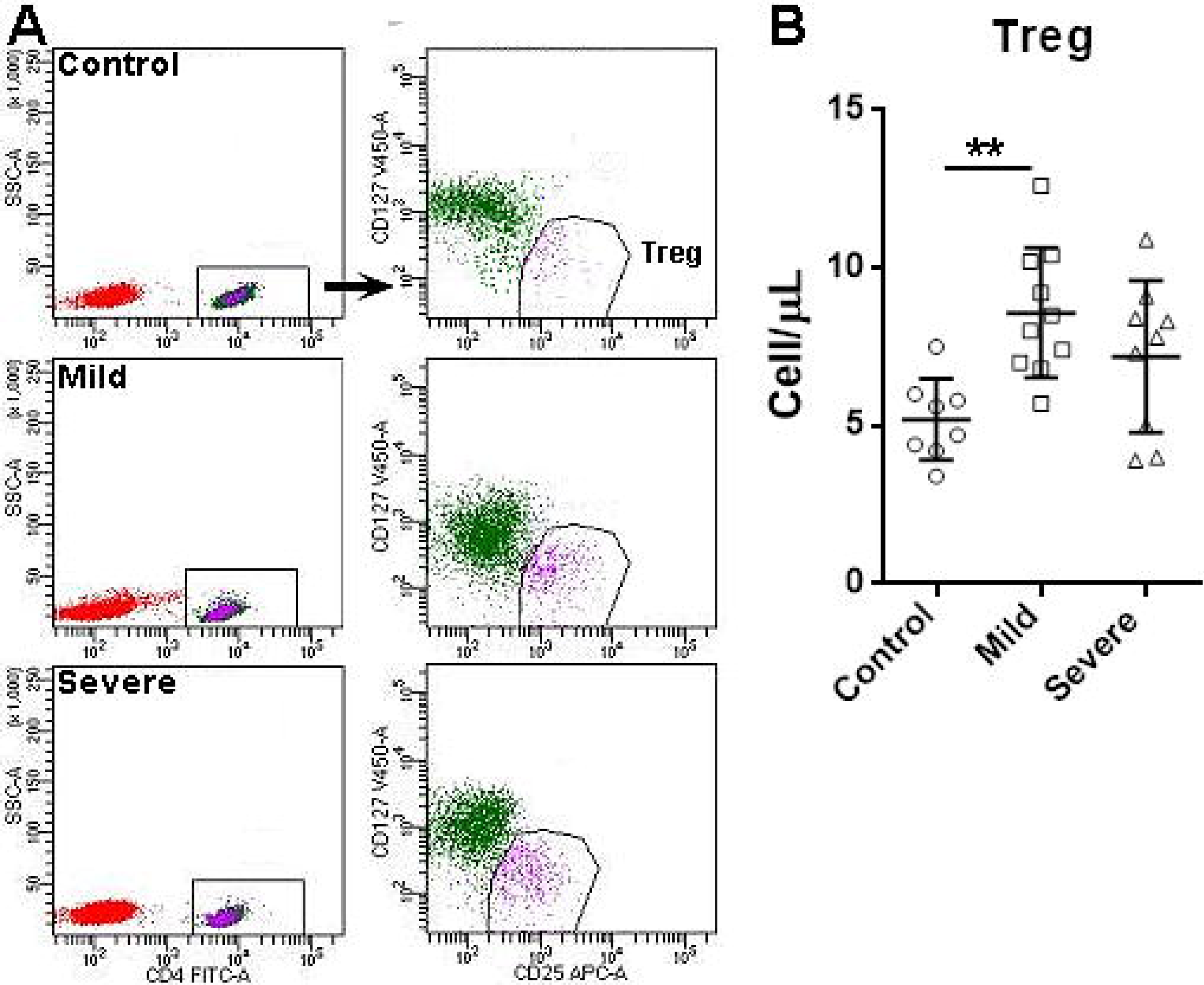
Cellularity of blood Tregs in COVID-19 patients at hospital admission. **(A)** Representative flow cytometry dot plots showing the gating strategy for Tregs. **(B)** Quantification of the number of Tregs. **, p<0.01.

### IL-2, IL-10, and IL-6 reflect the severity of COVID-19

Cytokines are crucial biomarkers of the progression of various inflammatory disorders including pneumonia. We characterized the expression of Th1 cytokine (IL-2 & IFN-γ), Th2 cytokines (IL-4 & IL-10), and pro-inflammatory cytokines (IL-6 & TNF-α) in the blood samples of patients at inpatient admission (Figure 4A). As demonstrated in Figure 4B, as compared with controls, IL-2 was considerably increased in severe patients but not in mild patients. IL-4 was remarkably raised in mild patients but not in severe patients (Figure 4C). IL-6 was significantly increased in severe patients but not in mild patients (Figure 4D). Like IL-2 and IL-6, IL-10 and TNF-α were significantly increased in severe patients but not in mild patients (Figure 4E & 4F). IFN-γ, however, showed no significant change in COVID-19 patients (Figure 4G).

**Figure 4.**
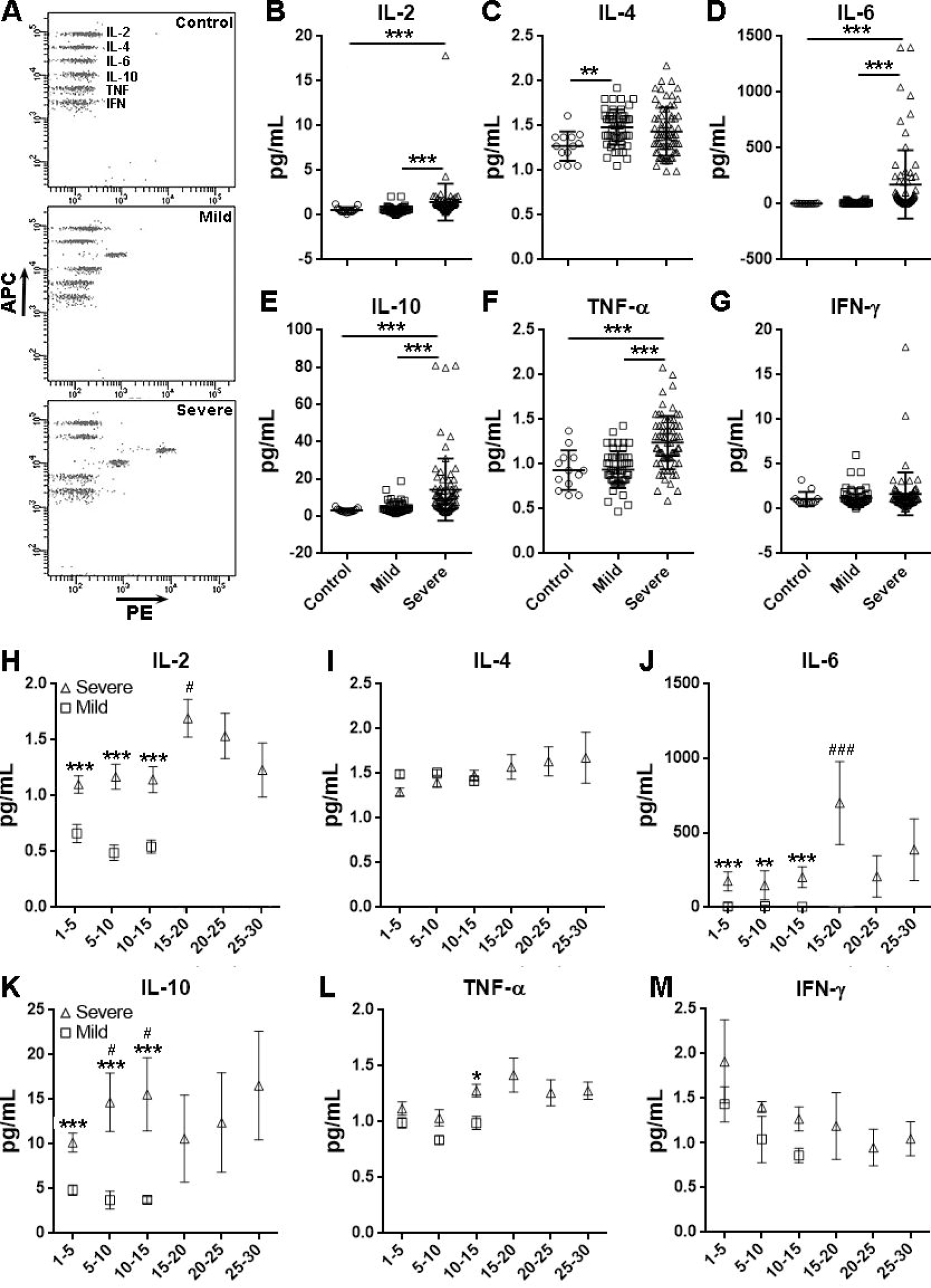
Cytokine levels in COVID-19 patients. **(A)** Representative flow cytometry dot plots showing the cytokine expression in patients. **(B to G)** Expression of indicated cytokines in COVID-19 patients at hospital admission. **, p<0.01; ***, p<0.001. **(H to M)** Expression of indicated cytokines in COVID-19 patients relative to the length of hospital stay. *, p<0.05; ***, p<0.001 in comparison to “Mild” of the same time interval. #, p<0.05; ###, p<0.001 in comparison to the corresponding “Severe 1-5” group.

We also evaluated the cytokine concentrations relative to the length of stay hospital. We found that severe patients always had a higher IL-2 level than mild patients in the same time intervals, and IL-2 reached a peak in severe patients on day 15-20, followed by a moderate decline. IL-2 was not significantly changed in mild patients along the stay (Figure 4H). No dramatic temporal change of IL-4 was observed in either mild patients or severe patients (Figure 4I). The IL-6 level in mild patients was always very low, while IL-6 was drastically increased in severe patients. Interestingly, as IL-2, IL-6 stayed at a relatively stable levels within two weeks, and then went up on day 15-20 followed by a decline back to the initial level (Figure 4J). IL-10, too, was always higher in severe patients than mild patients. Besides, IL-10 in severe patients underwent a further up-regulation on day 5-10 and day 10-15, respectively (Figure 4K). The changes in the levels of TNF-α and IFN-γ were almost neglectable (Figure 4L & 4M), suggesting that they probably are not good indicators of COVID-19 severity.

## Discussion

The outbreak of COVID-19 poses an urgent demand of understanding the role of immunity in the progression of viral infection and subsequent pneumonia. As a new coronavirus, SARS-CoV-2 is highly contagious probably owing to the virus spread through asymptomatic-infected individuals^6^. Until recently, very few publications characterized the comprehensive changes in the innate and adaptive immunity in SARS-CoV-2 infected people, although some non-peer reviewed sources such as Medrxiv released some information.

In the present clinical study, we analyzed almost all immune cell types in both mild and severe COVID-19 patients. We found lymphopenia consistent with former reports^4, 7, 8^. Neutrophils, however, were increased in severe patients. It is noteworthy that the lymphopenia seemed to arise from a comprehensive reduction of all lymphocyte populations, including CD4+ and CD8+ T cells, B cells, and NK cells. The decrease in blood lymphocytes might be due to the recruitment of reactive lymphocytes in the lungs. It will be helpful to investigate infiltrating cells in the lungs to test if the infiltrates are predominantly lymphocytes other than granulocytes. Another possibility is that lymphocytes are more sensitive than granulocytes to the disturbed homeostasis and then underwent significant cell death. Whatever the case, lymphopenia is an indicator of severe COVID-19.

Blood conventional T cells especially CD8+ T cells were reduced even in mild COVID-19 patients. This phenomenon might suggest the recruitment of reactive anti-viral CD8+ T cells into the infection site. According to reported T cell response in SARS-CoV infection, CD8+ T cell reaction was more frequent and robust than CD4+ T cell reaction^9^. It is therefore possible that this is also the case in COVID-19. Indeed, a latest case report shows the sign of excessive T cell activation in COVID-19, as evidenced by high proportions of HLA-DR (CD4 3.47%) and CD38 (CD8 39.4%) double-positive T cell fraction^5^. unpublished data indicate highly expanded clonal CD8+ T cells in the lung microenvironment of mild COVID-19 patients, suggesting a robust adaptive immune response connected to a better control of COVID-19 (https://www.medrxiv.org/content/10.1101/2020.02.23.20026690v1.full.pdf). However, B cell and NK cell number were also remarkably reduced in severe patients, suggesting a profound suppression of lymphocytes. Interestingly, Tregs, which play a crucial role in immunoregulation and anti-inflammatory response^10^, were elevated in mild patients, suggesting a possible immunosuppression. In severe patients, the Tregs cellularity was comparable to that in control individuals, probably due to an overall T cell deficiency. Although the reduction of lymphocytes is clear, the changes in the functions of lymphocyte populations remain unknown. In future it will be necessary to test the functions of distinct immune cell populations, so as to tell whether the innate or adaptive immunity is activated or impaired in COVID-19.

Recent research reveals the fatal cytokine storm in critical COVID-19 patients. Increased serum IL-6, IP-10, MCP-1, MIP-1A, and TNFα were observed in COVID-19 patients^8^. Several cytokine levels were also evaluated in our study. We found that serum IL-2, IL-6, IL-10 and TNF-α were mainly up-regulated in severe patients, while the first three cytokines exhibited a more meaningful increase. Thus, serum IL-2, IL-6, IL-10 could be used to assess the severity of COVID-19.

Cytokines are produced by both innate immune cells and adaptive immune cells. IL-2 is mainly secreted by activated T cells^11, 12^. The higher IL-2 level in COVID-19 patients likely reflects T cell activation. IL-6 is a well-known pro-inflammatory cytokine but it can also act as an anti-inflammatory mediator^13^. The cellular sources of IL-6 include myeloid cells, T cells, smooth muscle cells, endothelial cells, etc^14^. In COVID-19 patients, the dramatic increase in IL-6 likely originated from activated macrophages, consistent with another report^3^ and similar to the IL-6 up-regulation in SARS patients^15^. IL-10 acts as an anti-inflammatory cytokine deriving from alternatively activated macrophages, Th2 cells, Tregs, etc^16^. The significant increase of IL-10 in severe patients might be a negative feedback of the systemic and local inflammation. It is noteworthy that both IL-2 and IL-6 were dramatically up-regulated in severe patients on day 15-20 after inpatient admission and then declined. This similar pattern might suggest the aggravation of the disease and the following remission caused by therapies. However, more data are in demand to accurately explain the changes. Surprisingly, IFN-γ, which is a crucial anti-viral cytokine produced by both CD4+ T cells, CD8+T cells, NK cells and macrophages^17, 18^, remained at a low level in COVID-19 patients. It has been reported that IFN-γ participated in the cytokine storm in SARS patients^19^. Perhaps serum IFN-γ remains unchanged while IFN-γ in the lung tissue is elevated. More investigations are needed to test this hypothesis. Moreover, IFN-γ can downregulate the expression of the SARS coronavirus receptor ACE2 in a lab study^20^. SARS-CoV-2 seems to bind to the same receptor to infect target cells^21^. Hence, the potential effect of IFN-γ against SARS-CoV-2 infection needs further research.

In conclusion, our study shows that the comprehensive decrease of lymphocytes, and the elevation of IL-2 and IL-6 are reliable indicators of severe COVID-19.

## Data Availability

The author promised that all data are availability in the manuscript.

## Acknowledgement

Yaling Shi designed the study and wrote the paper. Mingkai Tan and Xing Chen collected samples and performed most immune cell detection. Yanxia Liu and Jide Huang conducted cytokine assays. Jingyi Ou did statistical analysis. Xilong deng evaluated Tregs.

The study is supported by Natural Science Foundation of Guangzhou (S2018010009732).

